# The Reputational Dividends of Collaborating with a Highly Reputable Agency: The Case of the FDA and Its Domestic Partner Agencies

**DOI:** 10.1101/2021.12.01.21267117

**Authors:** Moshe Maor, Raanan Sulitzeanu-Kenan, Meital Balmas

## Abstract

What, if any, dividends do agencies reap from collaboration with a highly reputable agency, such as the FDA? Utilizing a dataset covering 30 U.S. federal agencies over a period of 34 years (1980–2013), we estimate the short and long-term reputational effects of interagency collaboration. Collaboration is measured by the number of memorandums of understanding (MOUs) in effect between each agency and the FDA, while agency reputation is assessed using an automated measure of media-coverage valence (positive/negative tone) for each agency-year. To account for potential reverse and reciprocal causality, we utilize cross-lagged fixed-effects models. We find evidence of moderate rises in reputation due to increased collaboration with the FDA. These effects persist significantly for two years, before decaying to null after four years. Employing similar analyses, we furthermore estimate reversed causality – of reputation on the level of consequent collaboration – finding no evidence of such effects.

**Research Support:** Moshe Maor and Raanan Sulitzeanu-Kenan gratefully acknowledge financial support from the Israel Science Foundation under grant 1002/11.

## INTRODUCTION

While interagency collaboration is complex, with an abundance of risks and opportunities and no guarantee of a successful outcome (e.g., Ansell and Gash 2008; Bryson, Crosby, and Stone 2015; Finke 2020; Groenleer 2009; Mathieu et al. 2021; Wilson 1989), government agencies are increasingly expected to engage in collaborative relationships to solve pressing policy problems; reduce duplications, overlaps, and inconsistent regulation; increase the efficiency or effective use of resources between agencies; and address other ongoing challenges.^1^ It is anticipated that they will balance their turf- and autonomy-protecting tendencies against the potential benefits they can obtain by taking advantage of the programs, expertise, and resources possessed by other agencies that complement their core mission and that can, with effective collaboration, increase impact beyond what the agency can achieve alone.^2^ Consequently, government agencies are expected to have adequate mental, financial, and organizational capabilities to leverage these opportunities through a collaborative model that can significantly increase the effectiveness of all the partner agencies involved.

Most collaborations that come to mind in the U.S., for example, involve issues related to the core activities of the agencies involved, such as the collaboration between the Occupational Safety and Health Administration (OSHA) and the Environmental Protection Agency (EPA) on toxics; between the Food and Drug Administration (FDA) and the EPA regarding pesticide regulation; and between the U.S. Army Corps of Engineers and EPA concerning wetlands. The Endangered Species Act (ESA), which applies across all agencies and requires them to consult with the Fish and Wildlife Service (FWS) regarding impacts on listed endangered and threatened species, could be seen as a form of collaboration across putatively unrelated issues.

Existing research — particularly literature on collaborative governance, collaborative networks, governance networks, inter-organizational cooperation, and collaborative public management — has identified a variety of contexts and factors that determine the evolution of collaborations, the processes of collaboration, the formulation and negotiation of interagency collaborations, the governance and implementation of collaborations, as well as the substantial outcomes and evaluation of interagency collaborations (e.g., Ansell and Gash 2008; Bryson, Crosby, and Stone 2006; Emerson, Nabatchi, and Balogh 2012; Ward et al. 2018; Weible, Heikkila, and Pierce 2018). A few studies of interagency collaboration have furthermore focused on the concept of organizational reputation and the concept of perceived influence, also known as reputational power (e.g., Bernardo and Scholz 2010; Busuioc 2016; Fischer and Sciarini 2015; Gronow, Wagner, and Ylä-Anttila 2019; Ingold and Leifeld 2016; Knoke et al. 1996; Kriesi and Jegen 2001; Leifeld and Schneider 2012; Matti and Sandström 2013; Moynihan 2012). These studies demonstrate that inasmuch as interagency collaboration can advance agency reputation, it can also have negative effects in this regard, damaging reputational uniqueness (Busuioc 2016; Deruelle 2021), or have no effect whatsoever.

Surprisingly, little is known about the quantitative value of interagency collaboration for bureaucratic reputation. To capture the notion of reputational gains resulting from agency strategy, we coin the term *reputational dividends*, defined as the reputational value — measured by positive media coverage over an extended period — generated by a particular agency’s collaboration strategy. We use this term to posit a straightforward question: What reputational dividends, if any, do federal agencies reap from collaborating with a federal agency that enjoys a strong domestic and international reputation?

We endeavor to answer this question, discerning the short and long-term reputational effects of interagency collaboration, by examining a dataset encompassing 30 U.S. federal agencies over 34 years (1980–2013). Collaboration is measured by the number of memorandums of understanding (MOUs) in effect between each agency and the FDA.^3^ Agency reputation is assessed using an automated measure of media-coverage valence (positive/negative) for each agency-year. To account for potential reverse and reciprocal causality, we utilize cross-lagged fixed-effects models. We find evidence of moderate increases in reputation due to increased collaboration with the FDA. These effects persist significantly for two years, before decaying to null after four years. We employ similar analyses to estimate reversed causality – of reputation on the level of consequent collaboration – and find no evidence of such effects.

This paper breaks new ground in two ways. First, it extends scholarship concerning interagency collaboration methodology by relying upon Memorandums of Understanding (MOUs) over an extended period as a concrete marker of interagency collaboration. This is in stark contrast to much of the work conducted by scholars of political science (and other social sciences) vis-à-vis collaborative activities, which measures perceptions of collaboration via surveys and interviews. While some studies concerning public health have used MOUs to understand collaborative behavior (e.g., Khosla, Marsteller, and Holtgrave 2013), political science and public policy studies have not employed these kinds of more accurate measures.

Second, the paper extends our theoretical understanding of collaborative relationships and the potential reputational benefits that partner agencies may reap. It links scholarship concerning the critical role played by agency reputation as a determinant of agency regulatory behavior, extending this to posit that collaboration between federal agencies and an agency that enjoys a powerful reputation likely influences the resulting reputational dividends. This is important because as public problems become increasingly complex, interagency collaborations (as well as collaborative models involving the private and voluntary sectors) are expected to become progressively more important. A more nuanced understanding of the factors that enhance reputational dividends, which in turn may foster collaborative relationships with agencies that possess a strong reputation, is therefore necessary and constructive.

The remainder of the paper is structured as follows: first, we review the political science and network governance literature concerning the connection between government agencies’ collaboration and organizational reputation. Second, we present an analytical discussion that undergirds the hypothesis examined here. We then present descriptive results and the empirical findings. Finally, we discuss the practical and theoretical contributions of the paper, its caveats, and its deriving agenda for future research.

## RELEVANT LITERATURE

This study draws on developments in institutional political science that view regulators as generally rational agents (Zamir and Sulitzeanu-Kenan 2018) as well as politically conscious organizations interested in protecting their reputations. *Organizational reputation* refers to “a set of symbolic beliefs about the unique or separable capacities, roles, and obligations of an organization, where these beliefs are embedded in audience networks” (Carpenter 2010a, 45). If reputations are successfully formed, cultivated, and managed, they become “[…] valuable political assets – they can be used to generate public support, to achieve delegated autonomy and discretion from politicians, to protect the agency from political attack, and to recruit and retain valued employees” (Carpenter 2002, 491). As powerful assets, strong reputations can be deemed equivalent to agency coalition building (Carpenter 2001, 22), and various incentives encourage agencies to protect them. “There are other things that bureaucracies protect and ‘maximize’, but for many agencies […], reputation protection serves as the simplest and most powerful dynamic governing their behavior” (Carpenter 2004, 54). Consequently, numerous studies in this field are driven by the idea that “[…] when trying to account for a regulator’s behavior,” we should “*look at the audience*, and *look at the threats*” (Carpenter 2010b, 832; *italics in original*). Scholars claim that agency reputation-management strategies vary according to the number of reputational threats arising due to conflicting audience assessments vis-à-vis the agency’s end products, procedures, and performance (Carpenter 2010a; Carpenter and Krause 2012; 2015; Boon, Salomonsen, and Verhoest 2021; for a review, see Perez 2021). Potential sources of threats in risk regulation regimes (Hood, Rothstein, and Baldwin 2001; Rothstein 2003; Rothstein, Borraz, and Huber 2012) include the disparity between an agency’s policy conduct and reputation, low levels of organizational conduct (e.g., subunits fail to coordinate satisfactorily), and shifting expectations among the agency’s audiences regarding the agency or the sector within which it operates.

Recent findings have indeed related the ramifications of reputational concerns to the varying speed at which new drugs are approved (Carpenter 2002); the apportioning of funds to certain areas (Gilad 2012); the degree of an agency’s legal independence (Maor 2007); organizational task prioritization (Gilad 2015); endogenous construction of jurisdiction (Maor 2010); the extent to which regulatory errors are publicly observable (Maor 2011); the pace of regulatory enforcement (Maor and Sulitzeanu-Kenan 2013); a regulatory agency’s policy, regulatory, and scientific outputs (Krause and Douglas 2005; Maor and Sulitzeanu-Kenan 2016; Rimkuté 2018); cooperation outcomes (Busuioc 2016); regulatory enforcement (Etienne 2015; Gilad and Yogev 2012); democratic participation in an agency’s work (Moffitt 2010); accountability relations and behavior (Busuioc and Lodge 2016; 2017; Christensen and Lodge 2018); organizational politics (Blom-Hansen and Finke 2020), strategic communication (Anastasopoulos and Withford 2019; Baekkeskov 2017; Christensen and Lægreid 2015; Christensen et al. 2020; Christensen and Lægreid 2020; Gilad, Maor and Bin-Nun Bloom 2015; Grøn and Salomonsen 2019; Maor 2016a; 2020; Maor, Gilad, and Ben-Nun Bloom 2013; Moschella and Pinto 2019; Müller and Braun 2021; Rimkute 2020); and the initiation of procedures against EU member states (Veer 2021).

A few studies have focused on organizational reputation while analyzing interagency collaboration. For example, in the case of Hurricane Katrina, Moynihan (2012) found that “(i)f extra-network reputation and political responsibility is more important than intra-network reputation and norms of reciprocity, blame avoidance strategies are more likely” (p. 27). Furthermore, Busuioc (2016) demonstrated that strong turf-protective tendencies, and the lack thereof, are primarily shaped by the different reputational impacts of cooperation for the national authorities concerned: in some instances, cooperation with a European agency depletes national authorities’ important reputational resources; in others, it enhances national authorities’ ability to discharge their tasks successfully.

Research regarding collaborative governance and collaborative networks has also addressed how organizational reputation is formed in collective governance contexts. A stream of research revolving around collaborative disaster response demonstrated that the formation of organizational reputation is important for understanding actors’ behavior and network outcomes as produced by collaborative networks (e.g., Ansell, Boin, and Keller 2010; Christensen, Lægreid, and Rykkja 2016; Weick, Sutcliffe, and Obstfeld 2005). Another stream has demonstrated that the extent to which organizations are embedded in a network is associated with social outcomes in collaborative networks, such as reputation and influence, perceived salience, and trustworthiness (e.g., Chiu, Balkundi, and Weinberg 2017; Cooper, Scott, and Baggio 2009; Yang and Nowell 2021). Potential sources leading to these outcomes include, for example, the better access to information and social support enjoyed by centrally embedded organizations, which increases their autonomy and visibility in the network (e.g., Huang and Provan 2007; Ingold and Leifeld 2016; Provan, Hung, and Milward 2009). Another stream has evolved from the idea that forming ties with powerful network members can serve as a viable basis for power, thus focusing on whether an organization’s reputation in a network is explained by with whom it has ties (e.g., Agranoff 2007). Perhaps most important in this stream is the association found between organizations possessing certain characteristics (e.g., expertise, resources, unique roles) with a higher power in the network and subsequent increases in reputation in the field (Ran and Qi 2019). An evolving stream tries to disentangle the mechanisms through which network configurations explain organizations’ reputations. Yang and Lu (2021), for example, found that organizations’ *influence reputation —* i.e., the degree to which other actors regard a certain actor as influential and important in the policy process (Ingold and Leifeld 2016) — is explained both by how well organizations are connected in the network (degree of connectivity) and with whom they are connected (composition of ego networks).

Summing up, there are compelling reasons for scholars of bureaucratic politics to wish to delve into the linkages between bureaucratic reputations and interagency collaborations, rather than to focus their academic attention on collaboration’s outputs and outcomes, which may derive from these linkages. This leads us to hypothesize — in the following section — that government agencies collaborating with a highly reputable agency are likely to gain positive media coverage for a few consecutive years following the signing of a collaboration agreement.

## ANALYTICAL CONSIDERATIONS

An agency’s collaboration strategy and outcomes can exacerbate, moderate, or neutralize reputational threats, and, in some cases, even provide reputational dividends. To relate interagency collaboration and consequent reputational dividends theoretically, one must recognize that these dividends may derive from the way single agencies are perceived by multiple audiences and stakeholders — the context in which bureaucratic reputation is indeed often studied — as well as from the reputation collaborations possess. There are at least three substantial reasons for this. First, interagency collaborations are often matters of strategic choice (Lodge 2008) rather than a legal obligation, especially in the case of sister agencies within a federal department (e.g., the FDA and the National Institutes of Health [NIH] within the Department of Health and Human Services) as well as when the agencies involved in the collaboration recognize their responsibility and common objective to contribute to the total effort of the federal government in areas in which they possess recognized capability and expertise. Second, whatever the reason for the construction of a collaborative relationship, such relationships often involve considerable investments of resources (e.g., money, time) and expertise. And third, interagency collaboration is a repeated game with learning effects (e.g., Johnston et al. 2010), enabling trial-and-error, establishing trust and reciprocity, and allowing the envisioning of institutional change (Thomson and Perry 2006, 29). We therefore introduce the term *interagency collaboration reputation* and draw on signaling theory (Spence 2002; Connelly et al. 2011), as well as on Hood’s (2002; 2011) blame game approach, to explain the reputational dividends resulting from such a relationship.

An agency can use its collaboration strategy (read, choice of partners and of behavior during the collaboration) to signal to audiences and stakeholders information regarding one of the four dimensions of agency- and/or the collaboration’s reputation (Carpenter 2002), namely, the performative (does the agency/collaboration do its job?), moral (does the agency/collaboration protect the interests of its clients?), technical (does the agency/collaboration have the skills and capacity required?), and procedural (does the agency/collaboration follow accepted rules and norms?) dimensions. They can do so by more or less rationally choosing which dimension to stress vis-a-vis specific audiences. The literature describes this process as *prioritizing* (Maor 2015, 32), an act that all actors in the reputation game undertake simultaneously. Furthermore, each partner in interagency collaboration enters and leaves the collaborative relationship with its own expertise, capabilities, information, and, most relevant here, reputation.

Regarding the attribution of blame and credit, the higher the risk of blame following the collaboration’s actions, the higher the propensity for blame and thus for blame avoidance behavior, and vice versa for credit and credit claiming behavior. Some characteristics and outcomes of interagency collaborations may matter to heterogeneous audiences and stakeholders because they can affect their safety, well-being, and other aspects and dimensions of their lives or business operations (e.g., van der Veer 2021; Carpenter and Krause 2012; Etienne 2015; Maor 2016b; Boon, Salomonsen, and Verhoest 2021; Capelos 2016). Among the key characteristics that audiences may care about are perceptions of mutual understanding amongst partners involved in interagency collaboration; effectiveness of collaboration, particularly in areas related to the most serious threats to the public; intensity of interaction; flexibility of interaction; speed of learning; as well as perceptions of synergy among the activities of collaborating agencies. The perceived value of interagency collaborations (read, its reputation) is furthermore encompassed by the claim that the whole is worth more than the sum of its parts (Lewin 1951). Interagency collaborations, especially when subject to significant investment, can have a considerable impact on the perceptions and behaviors of audiences and stakeholders, and can therefore constitute both reputational threats and opportunities for the partners involved. Blame avoidance strategies (Hood 2011), which are designed to protect the reputation of the collaborating partners in cases of failure to address all areas of underlying sources of uncertainty, as well as regulatory crises and catastrophes, may be applied before, during, and after the collaboration.

Based on the discussion so far, we can envisage three sets of cases regarding the latter, some of which may be of relevance. The first set of cases involve interagency collaborations over a policy issue characterized by a relatively low level of salience. An example is the collaboration between the FDA and the U.S. Department of Agriculture, Science and Education Administration (SEA), formalized in an MOU signed in 1981 (46 FR 41859), concerning the use of drugs and feed additives in the diets of nonruminant animals. If low issue saliency remains unchanged throughout the collaboration, no reputational concerns emerge at the level of the agency and the collaboration concerned. For obvious reasons, such cases do not attract much academic attention.

The second set of cases encompasses interagency collaborations regarding a policy issue characterized by a relatively high level of salience, or which reaches high levels of salience during the collaboration. We refer here to public perceptions that a serious policy problem indeed exists and that the government could have possibly avoided or prevented this problem but failed to act appropriately. An example is food and dietary supplement marketers’ claims that their products are capable of preventing, treating, or curing various diseases, claims with no scientific basis, including, when appropriate, well-controlled human clinical studies. The reputational consequences of such collaborations, for both the partner agencies and for the collaboration itself, depend largely on the outcomes of the cooperation, as perceived by the multiple audiences of the collaboration itself and of each of the collaborating partners. Continuing with the above-mentioned example, the resulting collaboration between the FDA and the Federal Trade Commission (FTC) regarding food and dietary supplements during the Obama Administration revolutionized the regulatory landscape for these marketers. It involved FDA warning letters referencing potential violations of the FTC Act; FTC orders requiring that marketers gain FDA approval prior to using certain claims in advertising regulated by the FTC; and press releases announcing the simultaneous investigation of specific companies to assess violations of the Federal Food, Drug, and Cosmetic Act, enforced by the FDA, and the Federal Trade Commission Act, enforced by the FTC (Villafranco et al. 2011). An analysis of the perceived success of the collaboration, or its lack thereof, and the derived reputational implications often calls for a descriptive case-study analysis or an analysis of a small number of collaborations, and it is therefore beyond the remit of this paper.

Perhaps the most interesting set of cases involves interagency collaboration with an agency characterized by a strong domestic and international reputation. Such an agency may wish to enhance its regulatory activity by leveraging the impact of its actions through partnerships with other regulatory agencies, both domestically and internationally. A case in point is the collaboration between the FDA with FTC, Health Canada, and various state Attorneys General in “Operation Cure.All” concerning law enforcement and consumer education operations against internet health fraud in the early 2000s.^4^ Such inter-agency interactions may indirectly generate reputational threats for the collaboration and for its component agencies but may also offer them reputational advantages and even potential reputational dividends.

Regarding the threats, an agency that enjoys a strong reputation attracts considerable media attention, scrutinizing the agency’s collaborative strategy and actual collaboration relationships and outcomes. Once they enter the fray, partner agencies are therefore suddenly exposed to tight media scrutiny vis-a-vis the policy problem that the collaboration addresses, particularly in areas related to the most severe threats to the public. A highly visible collaborative relationship and outcomes can damage reputational uniqueness (Busuioc 2016; Deruelle 2021) and may also accidently spillover to actions unrelated to the collaboration, which otherwise would have remained below the media radar.

Furthermore, each partner agency can influence how multiple audiences perceive each one of the partner agencies. Critical actions carried out by partner agencies (e.g., warning letters, injunctions, recalls, and arrests) may attract media attention and significantly influence how other partners are perceived by multiple audiences and stakeholders. This, in turn, can have consequences for the salience of the policy problem that the collaboration addresses, and for the attribution of credit or blame (Weaver 1986; Hood 2011) among the collaboration partners throughout the cooperation and after its conclusion. However, when interagency collaboration faces challenging policy problems as well as crises, the powerful reputation of the renowned agency may moderate how severely the failure or crisis affects its partner agencies. In other words, an agency’s strong reputation may act as a buffer for how severely partner agencies suffer reputational loss when they are publicly accused of regulatory failures. Collaborating with an agency that enjoys a powerful domestic and international reputation may therefore provide not only reputational dividends but also ‘reputation insurance.’

The discussion so far brings our hypothesis into clear focus: *government agencies collaborating with a highly reputable agency are likely to gain positive media coverage for a few consecutive years following the signing of a collaboration agreement*. We therefore take advantage of the interest expressed by agencies that enjoy powerful reputations in collaborating with other agencies to further their core mission. By leveraging resources and expertise, and through cooperative agreements, agencies that enjoy a strong reputation can effectively collaborate to address critical public needs emerging within its jurisdiction and bridge scientific or professional gaps. This set of cases inevitably raises the potential of reverse causality – the impact of reputation (of partner agencies) on the likelihood of collaboration with a reputable agency. We will empirically address this issue here.

## RESEARCH DESIGN

To estimate the effect of collaboration with a reputable agency – the FDA – on the collaborating agency’s reputation, we compiled a novel panel-data based on the cooperation between and reputation of 30 U.S. federal agencies and departments over a period of 34 years (1980–2013). The dataset includes all agencies in the U.S. federal executive establishment that were operative during the period 1980–2013.^5^ The dataset was drawn from the *U.S. Government Manual* (USGM). Advisory, quasi-official, multi-lateral, educational, and support offices common to all cabinet departments were not included in the dataset. The agencies listed in the USGM were validated by a comparison with Lewis’s (2003) Administrative Agency Insulation Data Set, as well as with Lewis and Selin’s (2012) sourcebook of executive agencies. We consider an agency *terminated* if it was abolished or reorganized (as reorganization usually implies a change of the agency’s name).

### Collaboration with the FDA

We utilize the entire set of Memorandums of Understanding (MOUs) signed by the FDA and other agencies and government departments at the federal level to create a measure of cooperation with the FDA. This data was gathered by cross-referencing the following sources: (1) FDA database of domestic MOUs^6^; (2) the Federal Register (Notices, accessed via Lexis- Nexis database); (3) HeinOnLine database; and (4) Freedom of Information Act (FOIA) documents online. We excluded MOUs involving agencies operating at the state level, international MOUs (i.e., involving agencies overseas), and MOUs involving academic institutions. Overall, we identified 120 interagency MOUs involving a total of 535 partner agencies.^7^ These MOUs address collaborations in the following policy domains: agriculture; commerce; defense; education; energy; health and human services; homeland security; housing and urban development; interior; justice; labor; transportation; treasury and veteran affairs. The resulting measure of accumulated collaboration ranges between 0 (no MOU) and 28 accumulated MOUs with the FDA. Figure 1 presents the temporal variation in cumulative cooperation with the FDA during the research period for each of the 30 agencies and departments under examination. We employ the cross-sectional (between agencies) and temporal variation in collaboration. It is evident that some agencies did not sign a single MOU with the FDA [e.g., U.S. Office of Personal Management (OPM); U.S. Office of Special Counsel (OSC)], while others signed a sizable number of agreements, notably the NIH and EPA.

**FIGURE 1.**
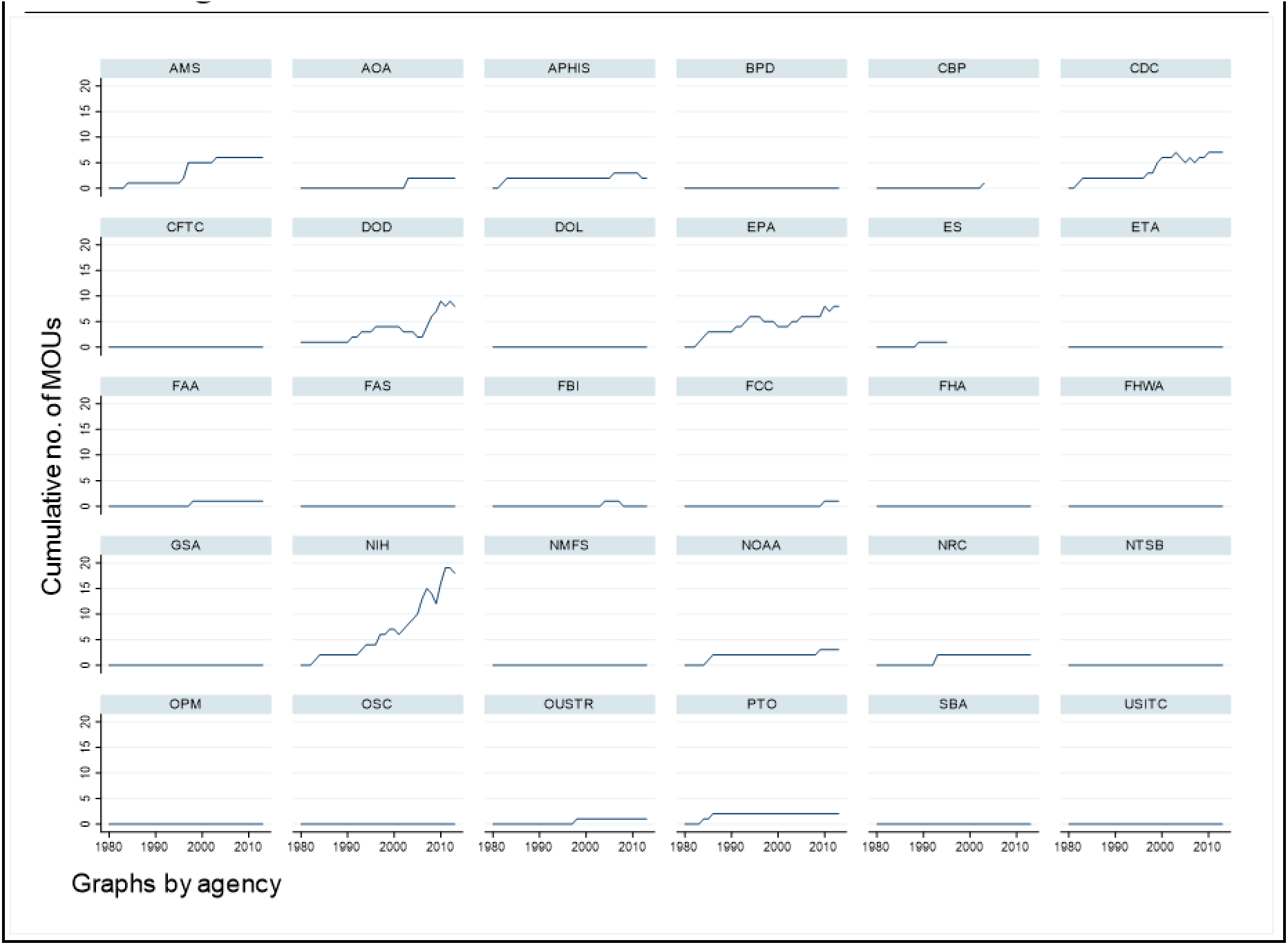
Temporal Variation in Cumulative Cooperation with the FDA Across Agencies, 1980-2013.

### Reputation data

Using automated content analysis, we obtained a measure of media valence for each agency- year, which constitutes our reputation measure.

#### The Media Sample and Method of Selecting News Articles

We analyzed the news coverage of the 30 U.S. federal agencies over a period of 34 years (1980– 2013). Three national newspapers were selected for the content analysis: *The Washington Post*, *The New York Times*, and *USA Today*. Those newspapers have the longest record on LexisNexis, dating from at least the early 1980s. Using LexisNexis, we collected all news articles that mention at least the name of one of the 30 agencies. For example, every news article mentioning the FCC in *The New York Times*, from 1982 until the end of 2013, was singled out. The complete database contains all news items from each of the above three newspapers that mention one or more of the agencies listed above (N=313,153). Three trained research assistants collected the data.

#### The media valence (positive / negative)

The large number of articles included in the content analysis performed for this study necessitated the use of computerized tools to capture the polarity of the dominant sentiment in each of the articles selected. The computerized content analysis tools employed were the Lexicoder Sentiment Dictionary (LSD) and the Amsterdam Content Analysis Tool (AmCAT), a web-based database and coding program.

The LSD, created by Young and Soroka (2012), is a comprehensive dictionary coded for valence. It is designed to target primarily the content of political news items but can also be used for other texts. Young and Soroka’s dictionary (2012) merges and standardizes three widely used and publicly available affective lexical resources from political science, linguistics, and psychology. The result is a broad lexicon scored for positive and negative tone, capturing the positive-negative sentiment valence in text. The LSD includes, for example, positive categories such as “benevolence,” “vindication,” “respect,” “cheerfulness,” and “intelligence,” as well as negative categories such as “insolence,” “malevolence,” “painfulness,” “disappointment” and “neglect.” Young and Soroka (2012) showed that the LSD produces assessments that are consistent with human coding (see also Soroka, Young, and Balmas 2015; Balmas 2017). Based on the LSD coding, it is reasonable to expect that, if a news article displays a higher frequency of negative as opposed to positive words, its general tone or context will be negative.

One of the goals of this research, however, is to capture the polarity of the tone a news article projects in covering a particular agency. To this end, the study employed AmCAT (van Atteveldt, Kleinnijenhuis, and Ruigrok 2008; Schultz et al. 2012), instructing it to search for words coded as either negative or positive by the LSD and occurring at a maximum distance of 10 words from the name of the agency. However, to conclude that a given negative or positive word refers to the agency itself rather than to a different entity, the results of AmCat must be compared with another, independently validated assessment – in the case in point, human coding. Such a reliability analysis was performed by three trained human coders and included 100 news articles.

Coders were instructed to read each article, identify the agency covered in it by the name, and then, for each agency in each article, designate the tone of coverage as positive, negative, or neutral. This validation required two reliability tests: one – between the three coders, and the other – the coders versus AmCat. The first test resulted in a Krippendorff’s Alpha of .71 and the second – .67. According to Hayes and Krippendorff (2007), with α=.70, the risk of accepting the data as reliable, when they are not, is quite low, q= 0.0125. This shows that, in most cases, when a negative or positive word appears at a short distance from another lexical item (signifying the agency), its tone will be associated with the entity signified. A staple concern in content analysis, whether human or automated, is producing reliable measures, relatively unaffected by random coding errors. One of the ways to deal with this issue is through volume. Aggregation works because errors are essentially random, thus canceling each other out as increasing amounts of data are added. Some texts may be coded a little too far in one direction or the other; by putting them all together, the correct signal is obtained. Dealing with large datasets requires automated content analysis, which, in turn, is usually performed on larger datasets.^8^

Although each of the 313,153 news articles was analyzed individually and coded as positive, negative, or neutral (the last, when the tone could not be identified), the unit of analysis employed in this study is not a single news article but rather an aggregated score for one year of coverage. The reason for the use of such a yearly measure is that the dependent variable, i.e., collaboration with the FDA, could only be gauged on a yearly basis.

The next stage, therefore, was to produce two measures that predict the probability of finding either positive (positive measure) or negative (negative measure) coverage of a certain agency in an article randomly chosen from a corpus of articles in which this agency is mentioned, selected in a single year. The negative measure was calculated by dividing the number of articles published over one year covering a given agency in a negative context by the number of all articles that mention that agency. The positive measure has the same denominator, but the numerator is the number of articles published over one year covering a given agency in a positive context. All news articles that were not identified as either positive or negative were categorized as neutral. Nevertheless, the yearly measures for positivity and negativity are not separated but combined into a single item. Therefore, our measure of reputation is the natural log of the ratio between the positive yearly valence rate and the negative yearly valence rate: 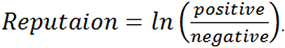.

Figure 2 presents the temporal variation in reputation during the research period for each of the 30 agencies and departments under examination.

**FIGURE 2.**
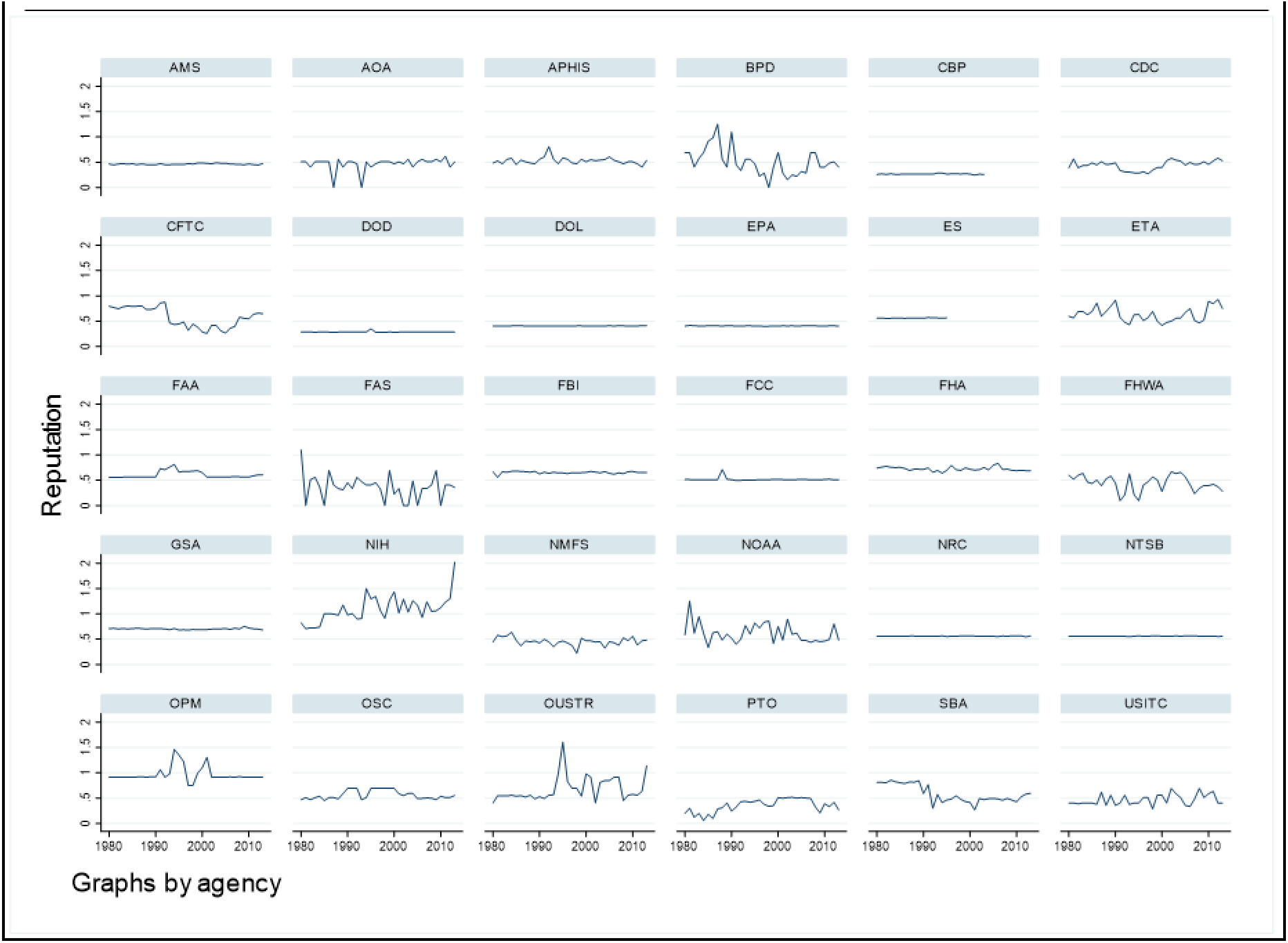
Temporal Variation in Reputation Across Agencies, 1980-2013.

### Analytic strategy

This study seeks to identify the effect of collaboration with a reputable agency – the FDA – on the collaborating agency’s reputation. Relying on the works of Carpenter (2001) and more generally Pierson (2011), which demonstrate the importance of disentangling causes and effects over time, our design allows for the possibility of reverse causality, namely that reputation may influence the propensity for collaboration. We employ cross-lagged panel models with fixed-effects (Allison, Williams, and Moral-Benito 2017; Moral-Benito 2013). The inclusion of agency fixed-effects enables us to control for time-invariant omitted variables. Reverse and reciprocal causality are addressed by including lagged values of the dependent variable – agency reputation – as predictors. This practice facilitates an estimation of the effect of the collaboration, even if it is influenced by pre-collaboration reputation level. Second, implementing a weaker sequential exogeneity assumption is required under conditions of potential reciprocal causality (Allison et al. 2017).

To map the duration of the causal effect of collaboration on reputation, we estimate a set of models varying from one to four-year lags. Given the potential bias due to mis-specified lags (Vaisey and Miles 2017), we follow recent advice to include the contemporaneous effect jointly with the lagged independent variable of interest (Leszczensky and Wolbring 2019; Nyberg et al. 2017). Our basic model is therefore:

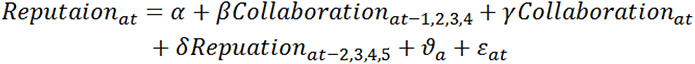

Where *Reputaion_at_* represents the reputation (measure of press valence) of agency *a* in year *t*, *Collaboration_at_*_-1_ is the measure of lagged cumulative collaboration with the FDA (number of MOUs in effect), and parameter *β* measures the average effect of collaboration on agency reputation. Lags of collaboration values vary between 1 to 4 years, across the four models. Alongside lagged values of collaboration, we include its current value, and lagged values of reputation – one year prior to the earliest lag of the collaboration, consistent with the causal ordering. The model includes agency fixed effects *ϑ_a_*. Note that although we have no information regarding each agency’s initial level of cooperation with the FDA prior to the research period (at the beginning of 1982), this variance among agencies is captured by the agency fixed-effects, allowing us to estimate the effect of changes in cooperation on agency reputation over the years. Analyses were conducted using the plm package in R, and all models are reported with robust standard errors.

## RESULTS

Table 1 presents four dynamic panel models with reputation as the dependent variable. Model 1 regresses our reputation measure on the contemporaneous level of collaboration, one-year lagged collaboration, and two-year lagged reputation. All models include agency fixed-effects. Model 2 differs only in that it includes the two- and one-year lagged collaboration and reputation, respectively. Models 3 and 4 follow the same method and use previous lagged measures of collaboration and reputation.

**TABLE 1.** Cross-lagged Fixed-effects Estimates of the Effect of Collaboration on Agency Reputation

|  |  |  |  |  |
| --- | --- | --- | --- | --- |
|  | Reputation |  |  |  |
|  | Model 1 | Model 2 | Model 3 | Model 4 |
| <u>Collaboration</u> |  |  |  |  |
| Current | -0.008<br>(0.007) | -0.005<br>(0.005) | 0.005<br>(0.004) | 0.012*<br>(0.006) |
| 1y-lagged | **0.024\***<br>**(0.011)** |  |  |  |
| 2y-lagged |  | **0.021\***<br>**(0.010)** |  |  |
| 3y-lagged |  |  | **0.011**<br>**(0.006)** |  |
| 4y-lagged |  |  |  | **-0.0002**<br>(0.006) |
| <u>Reputation</u> |  |  |  |  |
| 2y-lagged | 0.238***<br>(0.069) |  |  |  |
| 3y-lagged |  | 0.198**<br>(0.063) |  |  |
| 4y-lagged |  |  | 0.130<br>(0.067) |  |
| 5y-lagged |  |  |  | 0.137**<br>(0.046) |
| Agency FE | YES | YES | YES | YES |
| N | 932 | 902 | 872 | 842 |
| R-squared | 0.094 | 0.073 | 0.044 | 0.040 |
| Adj. R-squared | 0.062 | 0.039 | 0.008 | 0.002 |
| F Statistic | 31.025*** | 22.825*** | 12.954*** | 11.337*** |
| *Note: Robust standard errors in parentheses; \*\*\* p < .001; \*\* p < .01; \* p < .05* |  |  |  |  |

Model 1 shows that one-year lagged collaboration predicts an increase in reputation level. An increase of one MOU in the previous year is associated with a 0.024 increase in reputation – or 0.11 SD. This effect is identified when controlling for two-year lagged reputation and for contemporaneous collaboration. Note that despite the strong autocorrelation in the collaboration measure (one-year lagged collaboration accounts for 93% of the variance in current collaboration), we find no significant association between current collaboration and reputation. Model 2 provides similar (and slightly weaker) results for two-year lagged collaboration, suggesting that an increase in collaboration is associated with increased reputation for as long two years. Model 3 indicates a smaller positive yet statistically insignificant effect of 3-year lagged collaboration on agency reputation. Lastly, model 4 demonstrates a substantively null effect of 4-year lagged collaboration on reputation. Note that the coefficient for the current measure of collaboration in Model 4 is positive and statistically significant. However, given the results of Models 1 and 2 and the strong autocorrelation in collaboration, we can infer that this relationship is not causal but rather reflects the effects of lagged collaboration in the previous two years, omitted from this model. *These results suggest that an agency that increases its level of collaboration with the FDA should expect an increase in its reputation level in the two years following the collaboration*.

Figure 3 graphically presents the marginal effect of an increase of one MOU on the subsequent reputation of an agency, based on the analyses in Table 1. The figure shows significant positive effects of increases in collaboration that occurred one and two years before. The effect of a three-year old increase in collaboration is still positive, but smaller and statistically insignificant. The effect of a four-year old increase in collaboration is estimated to be null.

**FIGURE 3.**
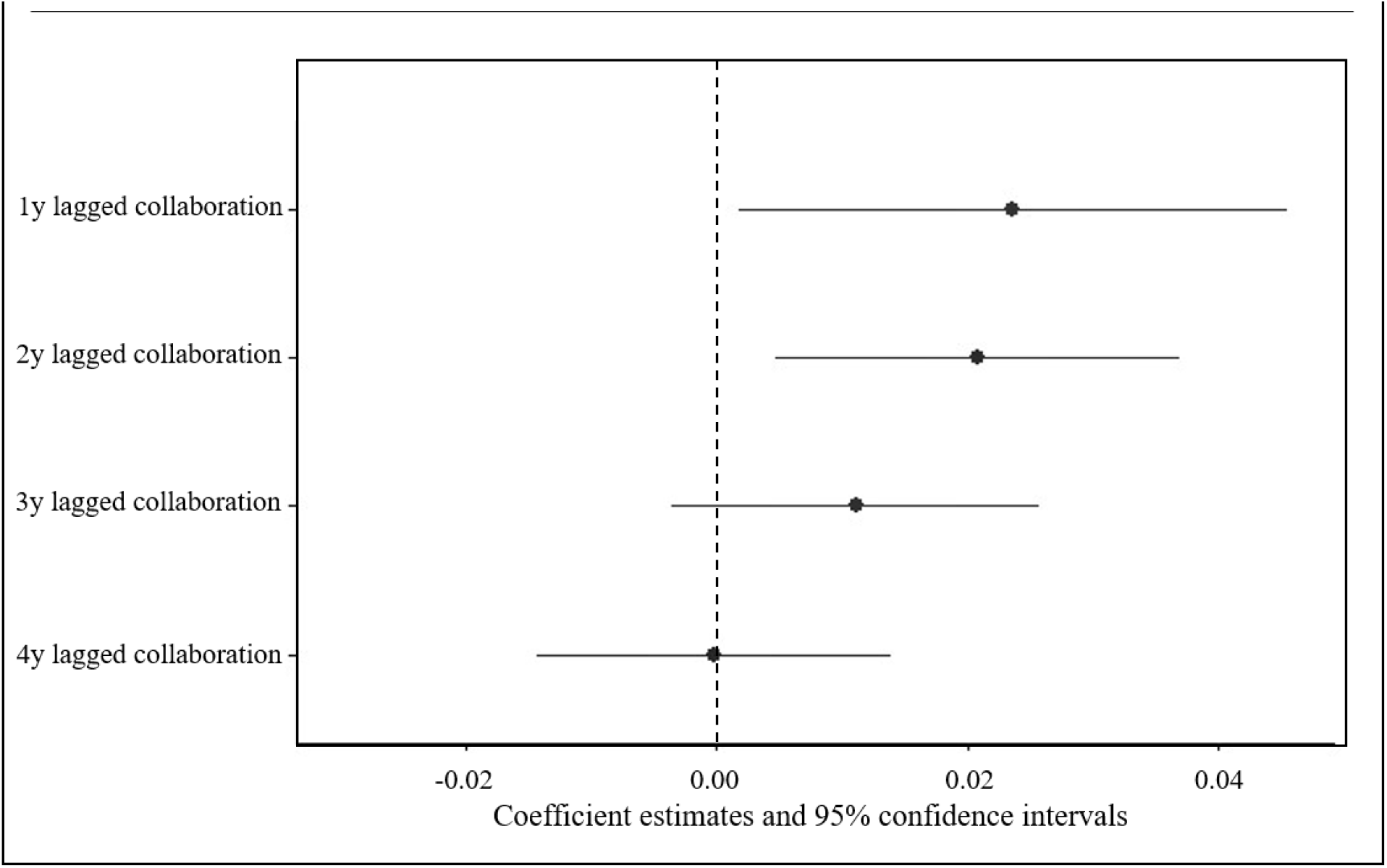
The Effects of Collaboration on Subsequent Agency Reputation

Although our interest is mainly in the effect of collaboration on agency reputation, we also consider the possibility of reciprocal causality. To address this, we conducted the same set of analyses by replacing the dependent variable with collaboration, using reputation as the main independent variable. Table 2 presents the results of these analyses. We find no evidence for either a lagged or contemporaneous effect of reputation on collaboration. These robust findings suggest that decisions on changes in the level of agency collaboration with the FDA do not appear to be influenced by reputation considerations. The theoretical implications of this combination of findings are elaborated in the concluding section.

**TABLE 2:** Cross-lagged Fixed-effects Estimates of the Effect of Collaboration on Agency Reputation

|  | Collaboration |  |  |  |
| --- | --- | --- | --- | --- |
|  | Model 5 | Model 6 | Model 7 | Model 8 |
| <u>Reputation</u> |  |  |  |  |
| Current | 0.016<br>(0.062) | 0.208<br>(0.140) | 0.436<br>(0.275) | 0.216<br>(0.200) |
| 1y-lagged | **-0.104**<br>**(0.084)** |  |  |  |
| 2y-lagged |  | **-0.147**<br>**(0.131)** |  |  |
| 3y-lagged |  |  | **0.005**<br>**(0.193)** |  |
| 4y-lagged |  |  |  | **0.129**<br>**(0.248)** |
| <u>Collaboration</u> |  |  |  |  |
| 2y-lagged | 0.982***<br>(0.046) |  |  |  |
| 3y-lagged |  | 0.985***<br>(0.094) |  |  |
| 4y-lagged |  |  | 0.991***<br>(0.136) |  |
| 5y-lagged |  |  |  | 0.969***<br>(0.164) |
| Agency FE | YES | YES | YES | YES |
| N | 932 | 902 | 872 | 842 |
| R-squared | 0.850 | 0.777 | 0.715 | 0.650 |
| Adj. R-squared | 0.844 | 0.769 | 0.704 | 0.636 |
| F Statistic | 1692.184*** | 1009.551*** | 701.677*** | 500.436*** |
| *Note:* Robust standard errors in parentheses; ***p < .001; **p < .01; *p < .05 |  |  |  |  |

## CONCLUSION

In this study, we unpack the reputational dividends reaped from interagency collaboration with a highly reputable agency. We define these dividends as the reputational value — measured by positive media coverage over an extended period — created by a particular agency’s collaboration strategy. We thereafter utilize a dataset covering 30 US federal agencies over the period of 34 years (1980–2013) to estimate the short and long-term reputational effects of interagency collaboration with the FDA. To account for potential reverse and reciprocal causality, we utilize cross-lagged fixed-effects models. We find evidence of moderate increases in reputation due to growing collaboration with the FDA. These effects persist significantly for two years, before decaying to null after four years. We employ similar analyses to estimate reversed causality – of reputation on the level of consequent collaboration – and find no evidence of such effects.

These findings have practical as well as theoretical significance. First, to the extent that the reputational dividends of collaboration with a reputable agency are expected, they provide a compelling incentive for reputation-sensitive agencies to collaborate with a highly reputable agency. In addition, if the agency’s decision to collaborate with a highly reputable agency is voluntary and not enforced by a central authority, then as far as reputation is concerned, the former agency has no incentive to defect from the collaborative relationship. Depending on the goal of the collaborative relationship, government officials and managers may want to build on the strength of their agency and seek repetitive formal collaboration with such an agency to enjoy these reputational fruits. Cognizant of this reputational value, it is plausible that elected executives concerned about their reputation will exert pressure on the agencies they control to collaborate with highly reputable agencies and thus reap reputational dividends. By the same token, it is highly probable that agencies less sensitive to their reputation will be more reluctant to collaborate with a highly reputable agency.

A notable null finding of this research is that the reputation of agencies is not predictive of their level of consequent collaboration with the FDA. This, together with the finding that such collaboration yields reputational dividends, can be accounted for in two different ways. First, the type of collaboration addressed by this study is not motivated by reputational considerations, even though it entails positive reputational dividends. Second, agencies possess reputation-based motivations to collaborate with the FDA, yet they are typically limited in their influence on the likelihood and extent of the collaboration, because either the FDA itself and/or other institutions are dominant in determining these outcomes. The latter institutions do not appear to consider the potential collaborating agency’s reputation in making such decisions.

This research is not without limitations. First, the extent to which the FDA is representative of federal agencies that enjoy a strong reputation may be questionable. Very few agencies worldwide enjoy such a powerful domestic and international reputation. Consequently, our findings may be atypical of other agencies that enjoy a strong reputation. Second, although MOUs provide a solid indicator of interagency collaboration, a sole focus on this formal form of collaboration implies that we are only looking at situations when this type of collaboration has occurred. Third, media content analysis for measuring organizational reputation suffers from some biases, such as negativity bias in the media. Nevertheless, since this bias is a constant feature, and our analysis utilizes the temporal variation in media valence, the effect of negativity bias in the context of this design is probably negligible. However, during the period under investigation, the newspaper industry saw remarkable changes in the scope and type of coverage. This may suggest a potential for measurement error when relying solely upon media coverage during this period.

Finally, our findings provide the baseline reputational outcome borne out by disentangling the reputation of the collaborating agency from other aspects of interagency collaboration, such as the duration of collaboration, the number of partners, the reasons for establishing the collaboration (e.g., legal requirement or voluntary), the nature of collaboration (e.g., joint decisions, information sharing), and so on. Future research should gauge the reputational dividends of each of these factors. Scholars may also address the question of whether, and under what conditions, the reputational dividends examined here can be obtained by the mere fact of collaborating with a highly reputable agency (i.e., by influencing perceptions) without solving the real policy problem targeted by the collaboration.

## Data Availability

All data produced are available online at

https://doi.org/10.7910/DVN/GX6AEU

## Footnotes

1 The term *collaborative relationship* refers to “any joint activity by two or more agencies, that is intended to increase public value by their working together rather than separately” (Bardach 1998, 8).

2 See, for example, Baldwin, Cave, and Lodge (2011, 368), Wilson (1989), and Busuioc (2016).

3 An MOU is defined here as a formal and non-binding inter-agency agreement that constitutes an understanding between the parties.

4 https://www.ftc.gov/news-events/press-releases/2001/06/operation-cureall-wages-new-battle-ongoing-war-against-internet

5 For the decision to use the term federal executive establishment rather than executive branch, see Lewis and Selin 2012, 1, note 1).

6 http://www.fda.gov/AboutFDA/PartnershipsCollaborations/MemorandaofUnderstandingMOUs/DomesticMOUs/default.htm

7 Each MOU represents a point in time at which agencies make collaboration decisions represented by each agency—MOU unit.

8 For the capacity of dictionary-based systems to capture tones of content, see, for example, Young and Soroka 2012; Soroka, Young, and Balmas 2015; Balmas 2017.

